# Using wastewater for population colorectal cancer screening and future research needs

**DOI:** 10.1101/2025.01.22.25320996

**Authors:** Elizabeth Wurtzler, Erica K Barnell, Catherine Morrison, Clayton Grass, Natalie DuPré, Donald J. Biddle, Allie Jin, Sandy Kavalukas, Rochelle Holm, Ted Smith

**Author notes:** Address correspondence to: Rochelle H. Holm, Center for Healthy Air, Water, and Soil, Christina Lee Brown Envirome Institute, School of Medicine, University of Louisville, 302 E. Muhammad Ali Blvd., Louisville, KY 40202 or Ted Smith, Center for Healthy Air, Water, and Soil, Christina Lee Brown Envirome Institute, School of Medicine, University of Louisville, 302 E. Muhammad Ali Blvd., Louisville, KY 40202. **Data Availability Statement:** Data generated in this study can be found in the published article and its supplementary files.

## Abstract

Colorectal cancer (CRC) is the third most common cancer and the second leading cause of cancer-related deaths in the United States. Individual screening is typically done with either a clinical stool-based test or direct clinical examination such as a colonoscopy. Given the low compliance with current screening recommendations and the high morbidity and mortality observed in areas with health disparities, we consider whether population-based testing using human RNA biomarkers in wastewater might effectively track the presence of CRC at the neighborhood level might be feasible. Wastewater samples were collected from four clusters in Louisville, KY: three representing cancer hotspots and one serving as a control neighborhood for feasibility data. Three wastewater replicates were obtained from each cluster. Human RNA biomarkers were isolated, quantified, and their RNA concentration levels were compared to clinical correlates. All replicates showed detectable levels of human cancer-associated RNA, including CDH1, which is a colorectal neoplasia-associated biomarker. Among CRC cluster sewershed samples, 8 of 9 replicate samples (89%) had a ratio of CDH1/GAPDH >=1 while the control sewershed sample showed ratio <1 for 2 of 3 samples. These preliminary data indicate that human RNA biomarkers can be detected in pooled community wastewater samples. While we have successfully identified the presence of these markers, further investigation with additional samples and closer alignment with documented case activity is necessary.

## Introduction

Prevention and early detection of colorectal cancer (CRC) is critical, as colon and rectal cancers have an estimated 152,810 new cases per year across the United States, making them the third most common cancer and the second leading cause of cancer-related deaths.^1^ CRC screening can be completed using an invasive procedure, such as colonoscopy, or an approved noninvasive test, such as the multitarget stool DNA test^2,3^ or the multitarget stool RNA test (mt-sRNA, ColoSense®)^4^. Fecal immunochemical tests (FIT), which measure hemoglobin levels in stool, are less sensitive and more specific than multi-target stool tests but are also used on an annual cadence for CRC screening.^5^ Population-based testing using human RNA biomarkers in wastewater might effectively track the presence of CRC at the neighborhood level and is an area which needs further research attention.

A large portion of new CRC cases in the United States present as late diagnoses due to increasing incidence in early onset CRC (individuals <50 years of age).^6^ Recently, screening guidelines recommended lowering the age of screening from age 50 to age 45.^7^ However, non-compliance with recommended guidelines, coupled with a rising incidence in individuals under 45, has led to colorectal cancer becoming a highly morbid disease among younger patients.^1,8^ Further, health disparities significantly contribute to the increased incidence of CRC and the diagnosis of later-stage disease with poorer outcomes.^9^ Factors such as unequal access to preventive care and screening, socioeconomic barriers, and disparities in health literacy disproportionately affect minoritized communities resulting in delayed diagnosis, less timely treatment, and a higher burden of advanced-stage CRC.^10^

Wastewater has been of remarkable value for infectious disease surveillance, but also has potential to monitor other population-level diseases.^11,12^ There is evidence from the COVID-19 pandemic that community level wastewater health data can be acted upon.^13^ Considering the existing CRC screening gaps, leveraging wastewater to detect elevated markers of colorectal cancer before clinical detection could help target areas to promote focused, individualized screening while also identifying regions with relatively lower likelihood of case discovery. Screening for CRC on a broader scale may offer a practical and cost-effective public health intervention to enable early diagnosis, particularly for communities experiencing barriers to cancer prevention and care.

Here we present feasibility data demonstrating the detection of colorectal neoplasia-associated RNA biomarkers in wastewater. These wastewater biomarkers adapt elements of the mt-sRNA test, which has shown high sensitivity and specificity for colorectal cancer and advanced adenomas in average-risk individuals over the age of 45. Employing conventional wastewater-based epidemiology methods, this study aimed to evaluate if RNA biomarkers used for the mt-sRNA test could feasibly be adapted to detect biomarkers of CRC through neighborhood-level wastewater samples.

## Materials and Methods

A retrospective study (The University of Louisville Institutional Review Board # 23.0319) at a tertiary care center analyzed CRC patients from 2021-2023. Residential addresses of these cancer patients^14^ and state cancer registry^15^ data were mapped to identify geographic clusters of high incidence rates of CRC (i.e., Cluster #1, #2 and #3) and their sewersheds were identified. An additional sewershed (Control #1) where none of the tertiary care patients lived was selected as a population control similar to CRC Cluster #2 based on similar population size, race distribution, and median household income (**Table 1**). All sewersheds were considered to be neighborhood-level sites. Population demographics were drawn from the ESRI Business Analyst data using the Enrich tool. No industrial wastewater flow is reported to have contributed upstream of any of the locations (**Table 1**).

**Table 1.** Information for the four clusters evaluated as part of this study, which included three colorectal cancer hotspots (CRC Cluster #1-#3) and one region to serve as a control area (Control Cluster #1). Total population associated with each sewershed area are provided. The distribution of race, ethnicity, and median household income, the number of tertiary care center patients, and state cancer registry patients for each sewershed area are also provided.

| **Sewershed** | **Region status** | **Total population** | **% Black** | **% White** | **% Hispanic** | **Median household income** | **Number of tertiary care center patients** | **Recent KY cancer registry patients** |
| --- | --- | --- | --- | --- | --- | --- | --- | --- |
| CRC Cluster #1<br>(MH04357) | Cancer Hotspot | 610 | 83.67% | 11.78% | 2.95% | \$19,202 | 4 | 2 |
| CRC Cluster #2<br>(MH08125) | Cancer Hotspot | 342 | 91.64% | 4.48% | 0% | \$50,000 | 1 | 2 |
| CRC Cluster #3<br>(MH03880) | Cancer Hotspot | 797 | 90.24% | 5.13% | 2.19% | \$37,204 | 1 | 0 |
| Control Cluster #1<br>(MH103715) | Control Area | 343 | 66.98% | 27.1% | 0% | \$67,919 | 0 | 0 |

Wastewater samples were collected three times over the course of the day via grab method from one street line utility hole in each sewershed on July 26, 2023 (N = 12). 175mL of influent wastewater was collected at each site at approximately 7:00, 10:00, and 13:00. Raw wastewater samples were stored at -20°C until analysis. 49 mL of each sample was analyzed. The mean of the triplicate samples over the course of the day were normalized.

Samples were blinded during quantitation assay runs. Prior to RNA extraction, the cellular material from each sample was pelleted using differential centrifugation.^16^ Samples were lysed and RNA was extracted using an EMAG (bioMerieux) by previously described methods.^16^ Extracted RNA concentrations were quantified using a Qubit. RNA expression values for GAPDH (housekeeping marker) and CDH1 (colorectal neoplasia-associated marker) were quantified via droplet digital PCR (ddPCR) using previously described methods.^4^

## Results

Total eukaryotic RNA was successfully extracted and isolated from all wastewater samples collected for the analysis. Average Qubit concentration across all samples was 13.8 ng / µL (range: 3.03 to 21.5 ng / µL) and average GAPDH concentration was 52.1 copies / µL (range: 0.45 to 202 copies / µL) (**Figure 1A-B, Supplementary Table 1**). Colorectal neoplasia-associated RNA biomarkers (CDH1), when normalized to housekeeping transcript (GAPDH), showed low concentration values for all clusters at 7:00 and 10:00, but showed a spike in value at 13:00 for CRC cluster #1 (**Figure 1C**). Average normalized colorectal neoplasia-associated RNA markers (CDH1 / GAPDH) for CRC clusters were 20.0 (CRC Cluster #1), 2.2 (CRC Cluster #2), and 4.0 (CRC Cluster #3); average normalized colorectal neoplasia-associated RNA markers (CDH1 / GAPDH) for the Control Cluster #1 were 2.6 (**Supplementary Table 1**). Interestingly, the CRC Cluster #1 had four times the number of known tertiary care center clinical cases than the other CRC Clusters #2 and #3 (**Supplementary Table 1**) and had higher average normalized CDH1/GAPDH than CRC Cluster #2 and #3. Additionally, the Control Cluster did not have any known tertiary care clinical cases at the time of this study and two of the three CDH1 / GAPDH measurements were less than one (**Supplementary Table 1**). Though the cases associated with the clusters were likely resolved at the time of wastewater collection, these areas have such elevated CRC incidence that it is more likely that undiagnosed cases are present here than in other locations.

**Figure 1.**
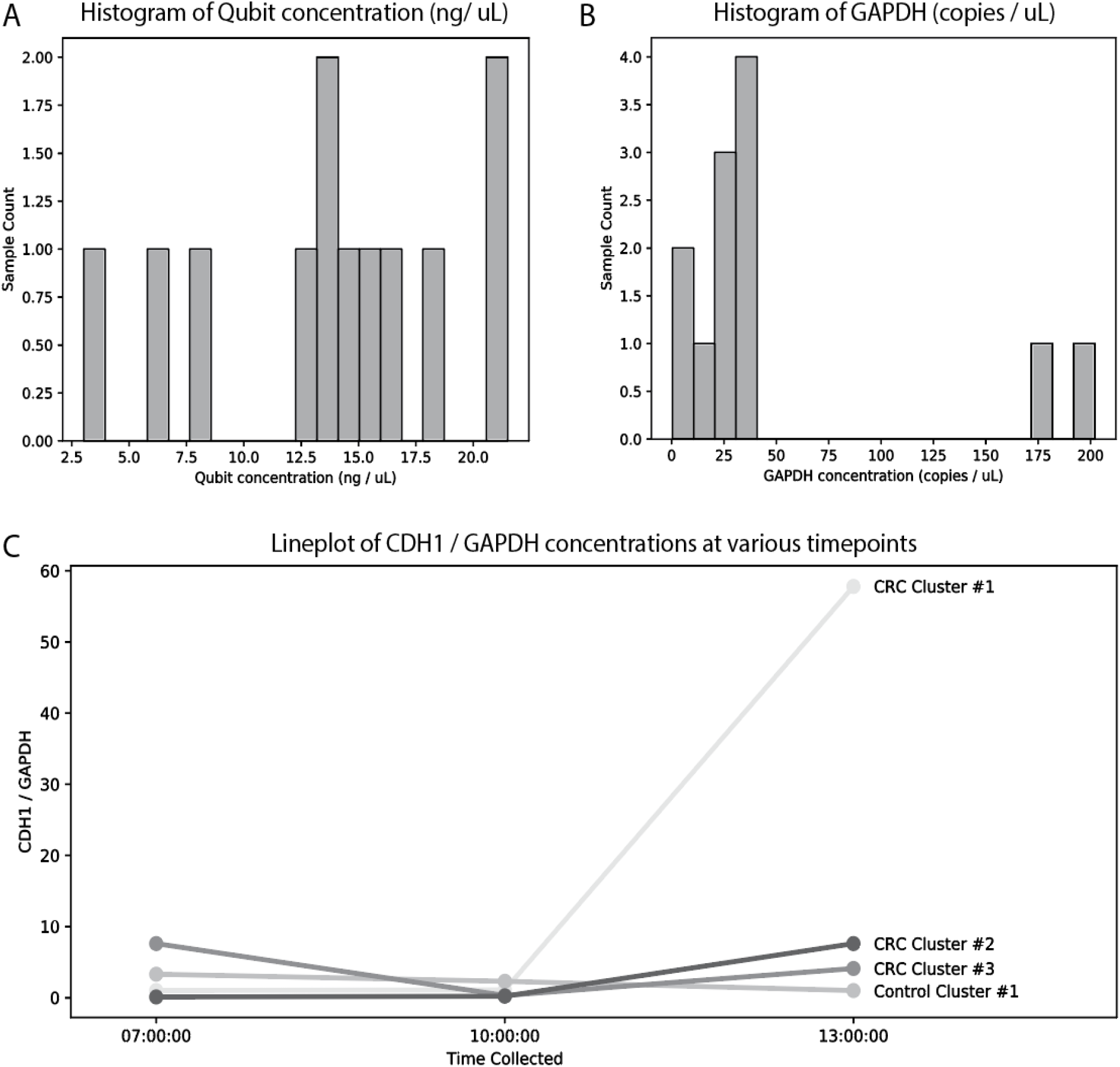
Assessment of RNA concentration values for all replicates across the four sewershed areas (CRC Cluster #1 - #3 and Control Cluster #1). **A)** Histogram of Qubit concentrations (ng / µL) for all samples assessed across the four sewershed areas. **B)** Histogram of GAPDH concentrations (copies / µL) for all samples assessed across the four sewershed areas. **C)** Lineplot of normalized colorectal neoplasia-associated RNA markers (CDH1 / GAPDH) for all samples assessed across the four sewershed areas parsed by various timepoints (7:00, 10:00, and 13:00).

All replicates from all four clusters showed detectable levels of human RNA, including CDH1, which is a colorectal neoplasia-associated biomarker.

## Discussion

The findings of this study underscore the potential feasibility of wastewater-based epidemiology to be expanded as a community population-level screening tool for colorectal cancer (CRC). By integrating environmental surveillance with biomolecular detection of CRC-associated markers, this study offers encouraging but not definitive preliminary data to help address the current limitations of CRC screening methodologies.

Previous research in this field has primarily focused on using wastewater to detect cancer treatment medications or environmental contaminants associated with cancer-related health outcomes.^17^ While some studies have explored the detection of mitochondrial DNA (mtDNA) mutations in wastewater, these mutations were not specific to a particular cancer type.^18, 19^ In contrast, the data presented here represent the first instance of using specific human RNA biomarkers for detecting colorectal cancer in wastewater.

The recent trend of increasing CRC incidence in younger populations and underserved communities underscores the need for improved public health approaches. While existing screening methods like colonoscopy and stool-based tests remain effective, their reliance on individual compliance presents challenges, particularly in communities facing socioeconomic barriers. While mail-based self-sampling has reduced the conventional visit-based CRC screening approach burden,^20^ wastewater surveillance could even further improve this screening access. Wastewater surveillance provides a complementary strategy to traditional screening by enabling a broader, community-level assessment of CRC risk without requiring individual participation.

Despite that our preliminary data justifies further study, this study has several limitations. Detection of colorectal neoplasia-associated biomarkers in the control area, where there are no known tertiary care center or KY cancer registry patients, needs further survey. The small sample size and geographic scope limit the generalizability of findings. Due to the retrospective nature, the timing of wastewater sampling and the number of known and active CRC cases precludes causal interpretations of the results and it remains unknown how the CDH1/GAPDH ratio in wastewater reflects relates specifically to CRC incidence. Additionally, the sensitivity and specificity of wastewater-based epidemiology for detecting CRC biomarkers need validation in larger populations.

Future research is needed in the following areas: 1.) Exploring the with more samples temporal, spatial and trend analysis; 2.) Developing a better understanding of signal integrity perhaps by tracing the wastewater of consenting CRC patients from the household plumbing source, neighborhood street line utility hole, and downstream treatment plant to determine sewer transport conditions and decay from patient source; 3.) Using cancer registry data associated with county-wide wastewater surveillance and to determine if CRC biomarkers detected in wastewater follow incidence gradients spatially; 4.) Widing the geographic distance between cluster and control groups to better determine the limitation of detecting clusters (based on the clinical boundaries); and 5.) Further development of intervention strategies that exploit CRC risk information from wastewater screening to increase the effectiveness of home-test or other targeted clinical screening programs.

This study demonstrates the earliest steps of feasibility testing have been taken for using wastewater analysis to detect CRC-associated RNA biomarkers. By combining environmental monitoring with targeted public health interventions, wastewater-based epidemiology holds promise to contribute to CRC prevention and early detection strategies, ultimately improving outcomes for high-risk communities.

## Data Availability

Data Availability Statement: Data generated in this study can be found in the published article and its supplementary files.

## Acknowledgements

We thank the Louisville/Jefferson County Metropolitan Sewer District for their valuable collaboration with the wastewater sample collection and Wayne Tuckson, Kevin Sokolowski, Adam Kaplin, David Hoetker, and Amy Moseley for project support and manuscript feedback.

## Author Contributions

Conceptualization: E.W., W.T., and T.S.; Methodology: E.W. and T.S.; Formal analysis: E.W., E.K.B, C.M., and C.G.; Writing-original draft preparation: E.W., R.H.H., and T.S.; Writing-review and editing: E.W., E.K.B, C.M., C.G., W.T., N.D., D.J.B., A.J., S.K., R.H.H., and T.S.; Supervision: E.W. and T.S. All authors have read and agreed to the published version of the manuscript.

## Financial Contributions

No external funding was received.

## Abbreviations

(CRC): colorectal cancer
(ddPCR): droplet digital PCR
(mt-sRNA): multitarget stool RNA

---

**Supplementary Table 1.** RNA concentration values for all replicates across the four sewershed areas (CRC Cluster #1 - #3 and Control Cluster #1). Region status, collection time, quality assessment (Qubit concentration (conc.)) and RNA concentration values (GAPDH and CDH1 copies / µL) are provided. Average normalized concentration values (CDH1 / GAPDH) have been calculated for each cluster.

| Sewershed | Region status | Time collected | Qubit conc | GAPDH copies / $\mu\text{L}$ | CDH1 copies / $\mu\text{L}$ | CDH1 / GAPDH | Average CDH1 / GAPDH |
| --- | --- | --- | --- | --- | --- | --- | --- |
| CRC Cluster #1 (MH04357) | Cancer Hotspot | 7:21 | 14.0 | 23.3 | 0.24 | 1.0 | 20.0 |
|  |  | 10:33 | 15.3 | 31.4 | 0.36 | 1.1 |  |
|  |  | 12:58 | 3.03 | 0.45 | 0.26 | 57.8 |  |
| CRC Cluster #2 (MH08125) | Cancer Hotspot | 7:30 | 18.6 | 6.3 | 0.21 | 3.3 | 2.2 |
|  |  | 10:30 | 16.6 | 23.9 | 0.54 | 2.3 |  |
|  |  | 12:40 | 20.8 | 29.4 | 0.29 | 1.0 |  |
| CRC Cluster #3 (MH03880) | Cancer Hotspot | 7:33 | 14.4 | 35.4 | 2.7 | 7.6 | 4.0 |
|  |  | 10:41 | 7.78 | 38 | 0.13 | 0.3 |  |
|  |  | 13:09 | 13.1 | 19.7 | 0.81 | 4.1 |  |
| Control<br>Cluster #1<br>(MH103715) | Control<br>Area | 7:45 | 21.5 | 180 | 4.2 | 0.1 | 2.6 |
|  |  | 10:20 | 6.63 | 202 | 0.38 | 0.2 |  |
|  |  | 13:00 | 13.6 | 35.7 | 2.7 | 7.6 |  |

